# Which traits predict elevated distress during the Covid-19 pandemic? Results from a large, longitudinal cohort study with psychiatric patients and healthy controls

**DOI:** 10.1101/2021.04.01.21254625

**Authors:** Katharina Brosch, Tina Meller, Julia-Katharina Pfarr, Frederike Stein, Simon Schmitt, Kai G. Ringwald, Lena Waltemate, Hannah Lemke, Katharina Thiel, Elisabeth Schrammen, Carina Hülsmann, Susanne Meinert, Katharina Dohm, Elisabeth J. Leehr, Nils Opel, Axel Krug, Udo Dannlowski, Igor Nenadić, Tilo Kircher

## Abstract

**Background:** The Covid-19 pandemic resulted in repeated, prolonged restrictions in daily life. Social distancing policies as well as health anxiety are thought to lead to mental health impairment. However, there is lack of longitudinal data identifying at-risk populations particularly vulnerable for elevated Covid-19-related distress.

**Methods:** We collected data of *N*=1268 participants (*n*=622 healthy controls (HC), and *n*=646 patients with major depression, bipolar disorder, schizophrenia or schizoaffective disorder) at baseline (2014-2018) and during the first lockdown in Germany (April-May 2020). We obtained information on Covid-19 restrictions (number and subjective impact of Covid-19 events), and Covid-19-related distress (i.e., subjective fear and isolation). Using multiple linear regression models including trait variables and individual Covid-19 impact, we sought to predict Covid-19-related distress.

**Results:** HC and patients reported similar numbers of Covid-19-related events, and similar subjective impact rating. They did not differ in Covid-19-related subjective fear. Patients reported significantly higher subjective isolation. 30.5% of patients reported worsened self-rated symptoms since the pandemic. Subjective fear in all participants was predicted by four variables: trait anxiety (STAI-T), conscientiousness (NEO-FFI), Covid-19 impact, and sex. Subjective isolation in HC was predicted by social support (FSozu), Covid-19 impact, age, and sex; in patients, it was predicted by social support and Covid-19 impact.

**Conclusion:** Our data shed light on differential effects of the pandemic in psychiatric patients and HC. They identify relevant, easy-to-obtain variables for risk profiles related to interindividual differences in Covid-19-related distress for direct translation into clinical practice.

## Introduction

The Covid-19 pandemic and its ramifications can be considered a global stressor (Kaye-Kauderer, Feingold, Feder, Southwick, & Charney, 2021). The impact of stressors on mental health depend on stressor characteristics, as well as traits of the exposed individuals (Alisic et al., 2014; Hensley & Varela, 2008; Kendler, Gatz, Gardner, & Pedersen, 2006; Marin et al., 2011). The Covid-19 pandemic has generated a qualitatively new challenge (Johns Hopkins University, https://coronavirus.jhu.edu/map.html).

Social distancing, a commonly implemented measure to limit virus spread, has put large parts of the population in social isolation for a prolonged period (Kira et al. 2020). This might also restrict availability of social support systems, including access to mental health care, leading to increased stressor load (Banerjee & Rai, 2020; Mancini, 2020). Social isolation is known to affect mental and physical health negatively, with effect sizes similar to premature mortality in hypertension and hyperglycaemia (Aleman & Sommer, 2020; Holt-Lunstad, Smith, Baker, Harris, & Stephenson, 2015; Holt-Lunstad, Smith, & Layton, 2010; J. Wang et al., 2017). Experts have warned that using social isolation as a strategy to contain the Covid-19 pandemic could result in a future mental-health pandemic, including increased suicide risk (“Keep mental health in mind,” 2020; Parrish, 2020; Reger, Stanley, & Joiner, 2020). In previous, smaller epidemics, such as severe acute respiratory syndrome (SARS), an increase in suicides was reported especially in elderly adults, which was associated with social isolation and anxiety (Yip, Cheung, Chau, & Law, 2010). First data from Japan show a 16% increase in suicides following the second wave of the Covid-19 pandemic (July-October 2020) (Tanaka & Okamoto, 2021).

Even though lockdown restrictions and infection rates vary across countries, current Covid-19 cross-sectional studies suggest elevated stress and negative affect increase across the general population worldwide. In a recent study in the Mexican population (*N*=1,105), half of the participants (50.3%) reported moderate to severe levels of psychological distress due to Covid-19, as well as 22.6% reporting moderate to severe symptoms of anxiety (Cortés-Álvarez, Piñeiro-Lamas, & Vuelvas-Olmos, 2020). In an American representative sample (*N*=10,368), more than 25% reported moderate to severe anxiety symptoms associated with the pandemic (Fitzpatrick, Harris, & Drawve, 2020). A review of four studies in the Chinese general population consistently found increased symptoms of anxiety and depression, as well as self-reported stress (Rajkumar, 2020). In another large cross-sectional study of the general Chinese population (*N*=56,679), participants reported several mental health symptoms, including depressive symptoms (27.9%), anxiety symptoms (31.6%), insomnia (29.2%), and acute stress symptoms (24.4%) (Shi et al., 2020). Reviewing 25 longitudinal studies and natural experiments in a meta-analysis, Prati and Mancini (2021) report a small, but highly heterogenous effect of lockdown measures on mental health across studies, and stress the need for further investigation of subgroups which might be at particular risk for adverse mental health effects.

Two meta-analyses reviewing mostly cross-sectional studies (up to May 2020) identified female gender, younger age (≤40 years), social isolation, and presence of chronic/psychiatric illness, among others, as risk factors for increased mental distress during the pandemic (Luo, Guo, Yu, & Wang, 2020; Xiong et al., 2020). Indeed, individuals with previous or current mental health problems constitute a vulnerable group, as (psychosocial) stressors are known to exacerbate existing symptoms of major depression (Burke, Davis, Otte, & Mohr, 2005; Kendler, Karkowski, & Prescott, 1999). The additional stress of the pandemic with concomitant social isolation might be particularly detrimental to individuals already at poor mental health. In a recent case-control study, psychiatric patients were shown to report higher worries about physical health, as well as higher anger and impulsivity symptoms compared to healthy controls during the pandemic (Hao et al., 2020).

While cross-sectional studies provide data of the impact on mental health across multiple populations, they lack longitudinal assessments that are better suited to examine the complex relationship between current and pre-pandemic mental health status. Indeed, Shanahan et al., (2020) showed that distress preceding the pandemic was the strongest predictor of emotional distress during the pandemic itself.

In this study, we analysed data on Covid-19 impact and subjective distress in an ongoing large longitudinal German bi-centre cohort, including patients with affective and psychotic disorders, and healthy individuals (Kircher et al., 2018). Participants of this unique large cohort had been deeply phenotyped, including structured diagnostic assessment, before the pandemic, and were now re-assessed after exposure to social distancing and isolation measures implemented during the lockdown in Germany. Trait markers, indicative of risk for mental distress or disorders, might modulate individual level of Covid-19 distress. Based on the literature, we selected eleven variables previously associated with mental health problems which are relatively stable across time, i.e.: childhood maltreatment, familial risk, social support, resilience, IQ, trait anxiety, and the Big Five personality traits (i.e. openness, conscientiousness, extraversion, agreeableness, neuroticism) (See Table 1). This allowed us to test 1) how Covid-19 restrictions impact HC and patients, and 2) which baseline variables predict Covid-19 distress.

**Table 1:**
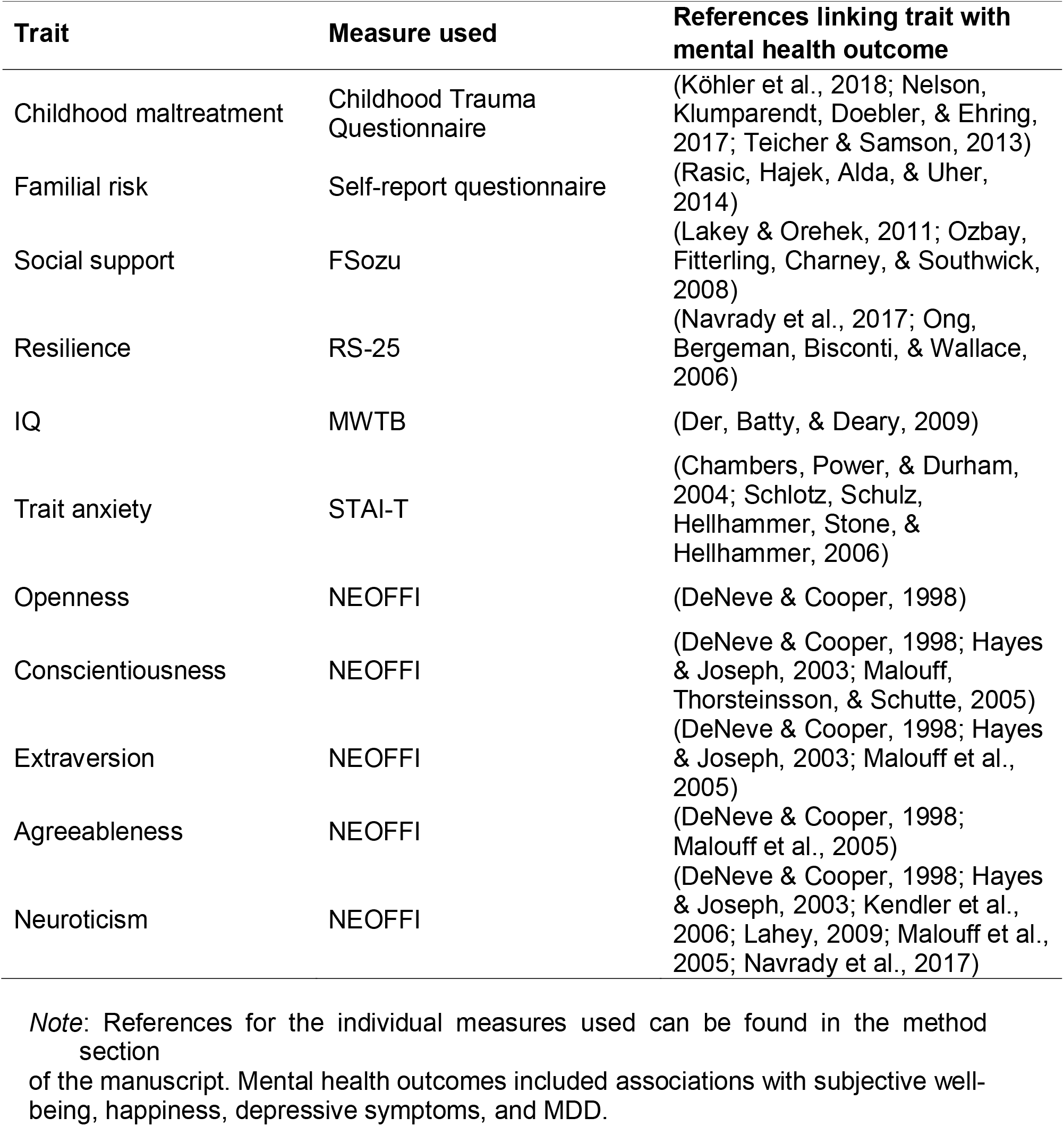
Baseline variables collected between 2014-2018 associated with mental health outcomes

## Method

### Sample description

The DFG FOR2107 cohort is a bi-centre, longitudinal study investigating healthy controls (HC), and patients with MDD, bipolar disorder (BP), schizophrenia (SZ), and schizoaffective disorder (SZA) (Kircher et al., 2018). The study was approved by the ethics committees of the universities of Marburg and Münster, Germany, in accordance with the Declaration of Helsinki. Baseline data used in this study were collected in Marburg and Münster from 2014 – 2018, with a median of data collected in 2016 (36.6%), four years before the Covid-19 telephone interview in 2020 was conducted.

From April – May 2020, during the first lockdown in Germany, we contacted *n*=1928 individuals who had previously participated in baseline testing. Of these, *n*=526 could not be contacted, and *n*=134 did not want to participate in the telephone survey, thus leaving a final sample of *N*=1268 (65.5% female; HC *n*=622, patients *n*=646; MDD *n*=514, BD *n*=74, SZ *n*=33, SZA *n*=25). Based on the literature, we chose eleven traits associated with mental health outcomes: Big Five (NEO-FFI), social support (FSozu), IQ (MWT-B), childhood maltreatment (CTQ), familial risk (assessed using a questionnaire), resilience (RS-25), and trait anxiety (STAI-T) (Costa & McCrae, 1992; Fydrich, Sommer, & Braehler, 2007; Laux, Glanzmann, Schaffner, & Spielberger, 1981; Lehrl, 2005; Leppert, Koch, Brähler, & Strauß, 2008; Wingenfeld et al., 2010). Table 2 lists sociodemographic and test data at baseline.

**Table 2:**
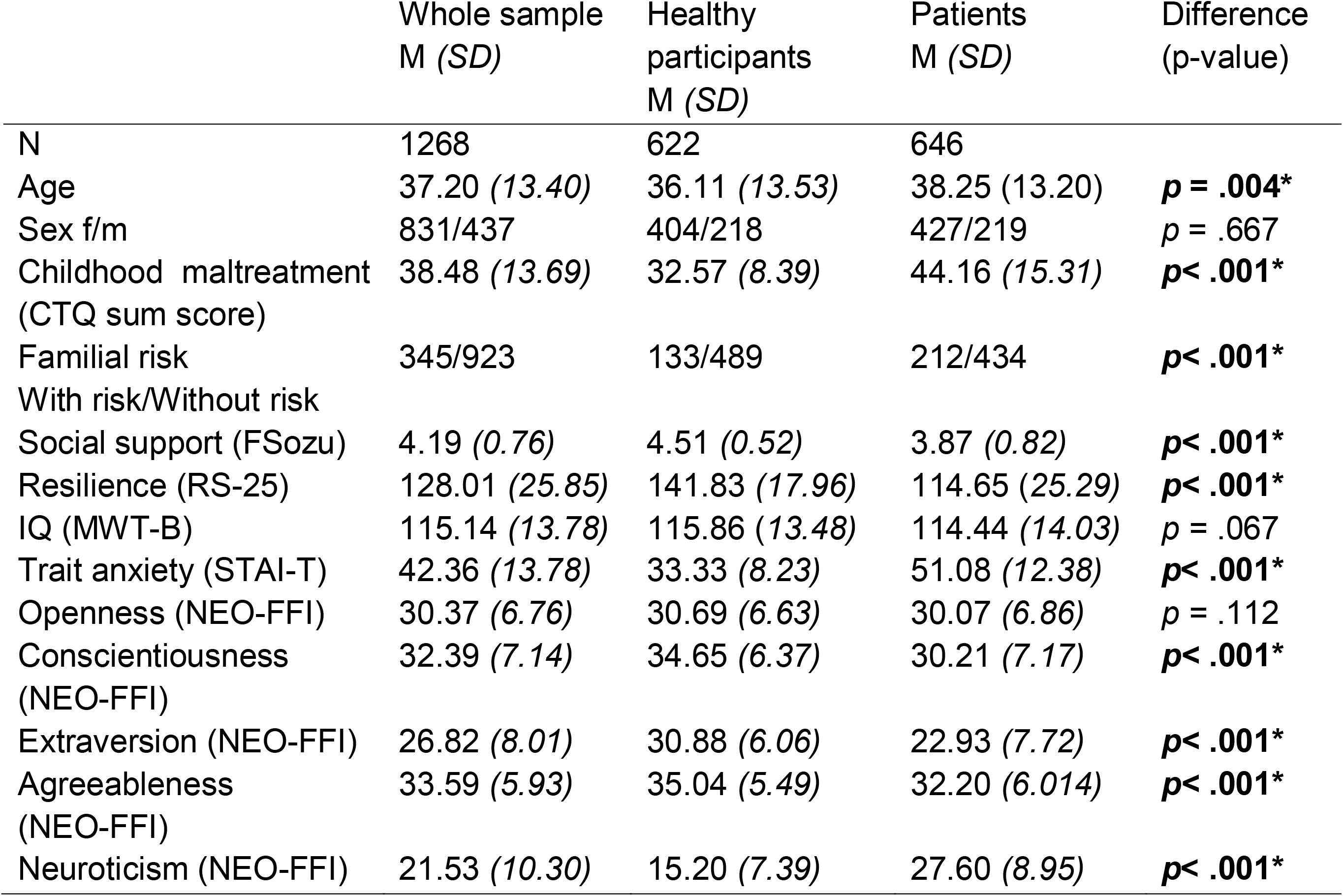
Sample characteristics at baseline collected 2014-2018 before the COVID-19 pandemic

### Data collection

Social distancing measures and formal regulations issued by the German Federal Government on March 22^nd^, 2020 included severe reductions of interpersonal contacts in both social (e.g., closing of gathering places, restrictions in meeting people) and occupational areas of daily life (e.g., closing of schools, working from home). The restrictions were in place during the entire data collection process.

We obtained lockdown data from April 7^th^, 2020 – May 8^th^, 2020. Trained research assistants conducted 20-minute semi-structured telephone interviews. The team was briefed to provide information about help hotlines if participants reported increased psychiatric symptoms due to Covid-19. We assessed *Covid-19 impact* through self-report of Covid-19-related incidents (Table 3). *Covid-19 distress* was operationalized as Covid-19-related subjective fear and isolation (Table 4). Covid-19-related incidents included 11 types of events, i.e. quarantine, working from home, loss of recreational activities, cancellation of private travel, cancellation of work travel, loss of childcare options, paused studies at school or university, restrictions in health care, pecuniary damage, short-time work, and loss of job. If participants had experienced such an incident, they were asked to rate its valence (positive or negative) and intensity (0 – 5 Likert-scales, with higher scores indicating higher intensity). *Covid-19 impact* was calculated by adding all intensity-related ratings, irrespective of their perceived valence. *Negative Covid-19 impact rating* and *positive Covid-19 impact rating* were calculated by only adding intensity ratings for either negatively, or positively rated items, respectively. This was based on scoring done in the life event questionnaire (LEQ) (Norbeck, 1984). In addition, all participants were asked to give information regarding their work situation (type of employment) in January 2020, before restrictions were implemented in Germany.

**Table 3:**
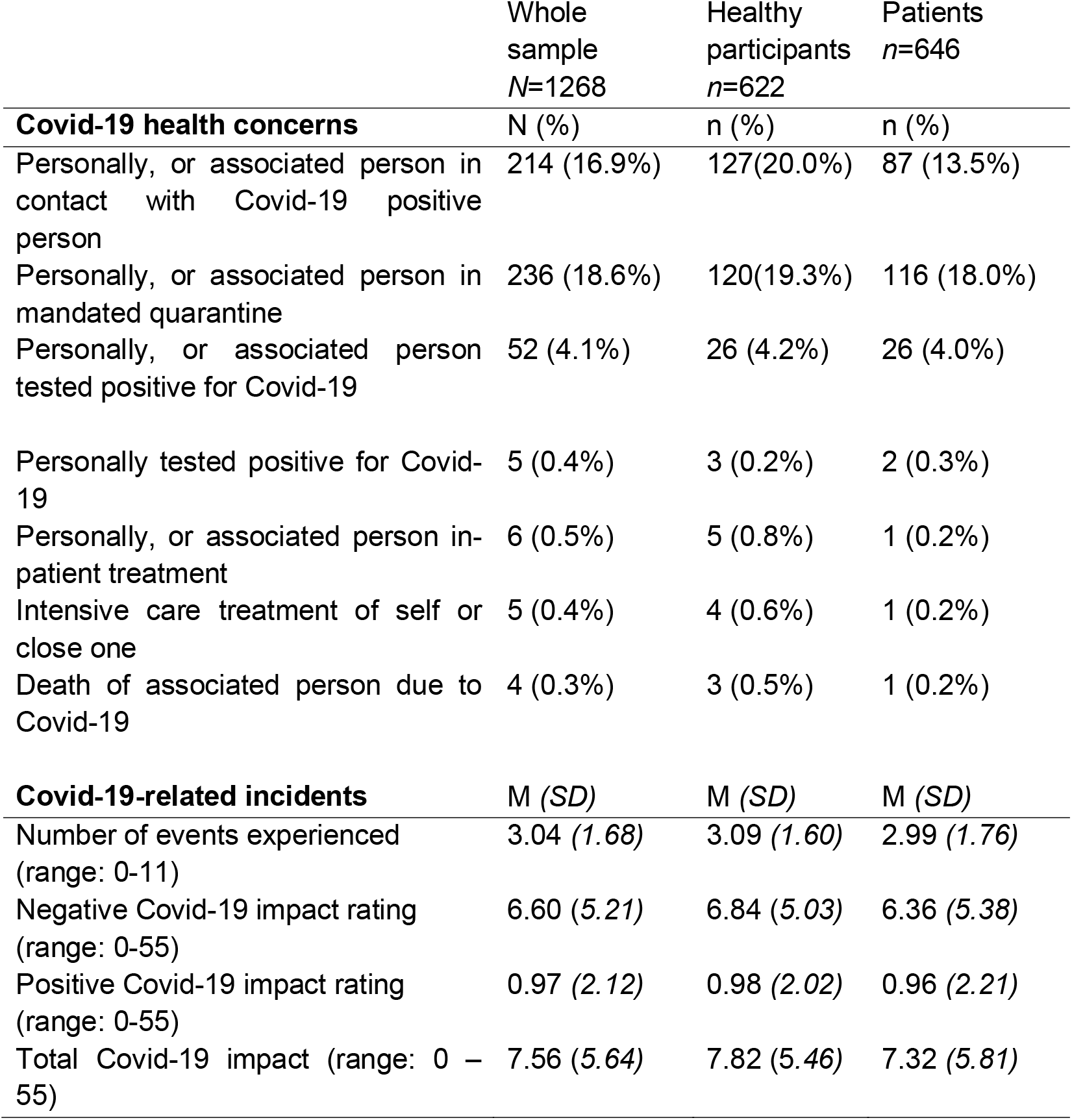
Health and life changes due to Covid-19. Data collected April/May 2020 during lockdown.

**Table 4:** Subjective Covid-19-related distress

|  | Whole sample | HC | Patients | p-Value |
| --- | --- | --- | --- | --- |
| Subjective fear (range 0-30) | 13.18 (5.27) | 12.96 (4.89) | 13.39(5.60) | *p* =.152 |
| Subjective isolation (range: 3-12) | 5.34 (2.20) | 4.73 (1.77) | 5.93 (2.41) | ***p*<.001**<br>* |

To better understand the impact of Covid-19 in the domain of health, we interviewed participants whether they themselves or associated persons (partner, core family, or friends) were subject to one or more of the following health events: testing positive for Covid-19, experiencing quarantine, contact to a Covid-19 positive person, any in-patient treatment, intensive-care treatment, or death due to Covid-19.

To assess participants’ subjective distress during the pandemic, we obtained self-report ratings of Covid-19-related subjective fear (subjective fear) and Covid-19-related subjective isolation (subjective isolation). Subjective fear was calculated by adding the ratings of three items “fear of Covid-19 regarding own health”, “fear of Covid-19 regarding health of associated persons”, and “fear of Covid-19-related economic and social consequences”, each rated on a six-point-Likert scale. Subjective isolation was assessed by prompting participants to think about current lockdown restrictions and then to answer three items of the revised UCLA loneliness scale (“There is no one I can turn to”, “I feel left out”, and “I feel isolated from others”), rated on a four-point-Likert scale (Russell, Peplau, & Cutrona, 1980). Subjective isolation was calculated by summing up the three items. Cronbach’s alpha for this scale was *α*=.684.

Patients were additionally asked to rate subjective changes in psychopathological symptom severity since the beginning of the pandemic (improved, unchanged, a little worse, substantially worse) as well as constraints regarding access to psychiatric/psychological care (see Table 5).

**Table 5:**
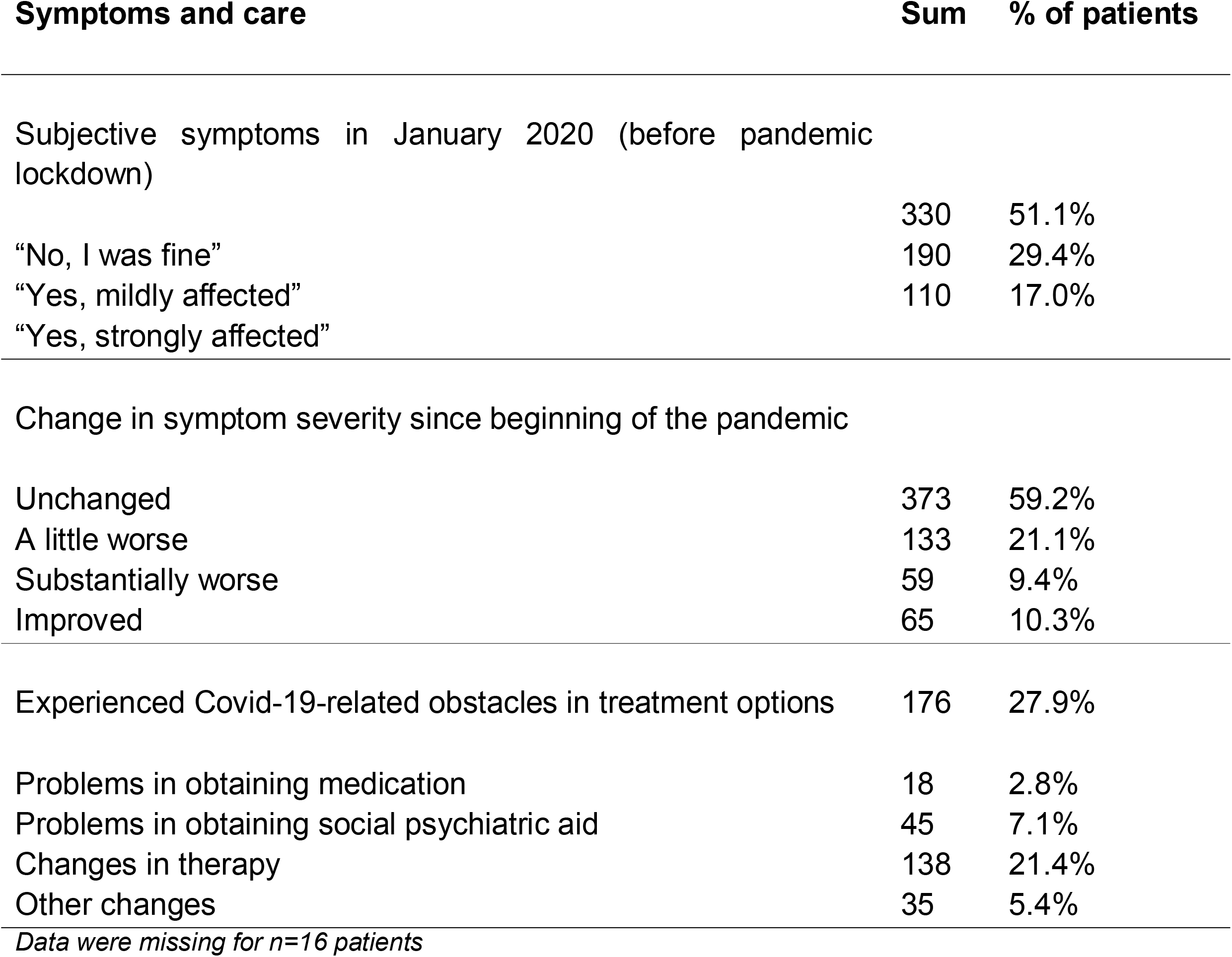
Changes in illness severity and treatment obstacles during pandemic in patients. Data collected April/May 2020 during lockdown.

### Statistical analyses

Significance level was set at α<.05. Analyses were performed using SPSS 27 α statistical software (IBM Corp.). Figures were designed using Microsoft Excel (Version 16.46) and ggplot2 running under R (Version 4.0.3) (Microsoft Corporation, 2018; R Core Team, 2020; Wickham, 2018). To compare mean scores between patients and healthy controls, two-tailed, independent sample t-tests were used. As Levene’s test for equality of variances was found to be violated for subjective isolation and fear, we report t-statistics not assuming homogeneity of variance for these. Frequency distributions were analysed using chi-square tests. As baseline data on remission status were missing for *n*=42 patients, and symptom change data were missing for *n*=16, we report the number of data sets used for each analysis. Patients with unchanged, improved, and worsened symptoms were compared using ANOVA.

Multiple regression analyses were applied to predict Covid-19 distress (i.e. Covid-19-related fear and isolation) from the eleven baseline variables, age, sex, site, and Covid-19 impact. Subjective fear was additionally predicted using diagnosis. We predicted subjective fear in the entire sample, while we ran multiple regression analyses for subjective isolation in HC and patients separately (as they differed significantly in this measure). For the multiple regressions, we performed Bonferroni correction for multiple testing, adjusting *p-*values for three tests. No multicollinearity was present (see Supplement, Table 1-3).

## Results

### Impact of Covid-19 restrictions on HC and patients

18.6% of the participants reported mandated quarantine either for themselves, or of an associated person (partner, family member or friend). Regarding Covid-19-related incidents, only 36 participants (2.9%) did not experience any event, the median of Covid-19 incidents experienced was 3. Patients and HC did not differ in the number of Covid-19 incidents experienced (*t*(1239)=1.02, *p*=.308, *d*=1.68), nor in their positive Covid-19 impact rating (*t*(1266)=0.162, *p*=.871, *d*=2.12), or negative Covid-19 impact rating (*t*(1266)=1.657, *p*=.097, *d*=5.21), and neither in their total Covid-19 impact rating (*t*(1266)=1.591, *p*=.109, *d*=5.64).

Regarding Covid-19 distress, patients did not differ significantly in subjective fear compared to HC (*t*(1254.22)=-1.44, *p*=.152, *d*=5.27). However, patients reported significantly higher subjective isolation, *t*(1183.57)=-10.16, *p<*.001; *d*=2.12.

27.9% of patients reported Covid-19-related problems in obtaining treatment options, of which 21.4% reported changes in therapy (Table 5).

Half of patients (51.1%) in our sample reported no psychiatric symptoms directly before the beginning of the pandemic. However, almost one third of patients (*n*=192, 30.5%) reported small to substantial symptom aggravation since the beginning of the pandemic, while 10.3% of the patient sample reported improved symptoms, and more than half of the sample (59.2%) reported no change in symptom severity (Table 5).

Based on changes in symptom severity, patients were split into three subgroups with unchanged, improved, or worsened symptom load (See Figure 2, and Table 6).

**Table 6:** Patients grouped according to symptom changes since the pandemic.

| Changes in symptom severity from January to April/May 2020 | Unchanged | Worsened | Improved | difference (p-value) |
| --- | --- | --- | --- | --- |
| Total n | 373 | 192 | 65 |  |
| **Baseline data** |  |  |  |  |
| Sex (f/m) | 226/147 | 143/49 | 49/16 | **p=.001\*** |
| Trait anxiety | 49.95 (12.87) | 53.82 (10.67) | 50.27 (12.88) | **p=.002** |
| Conscientiousness | 30.68 (6.96) | 29.53 (7.61) | 29.51 (7.32) | **p=.148** |
| Social support | 3.89 (0.84) | 3.76 (0.81) | 4.02 (0.69) | **p=.054** |
| **Diagnosis at baseline n** |  |  |  | **p=.755** |
| MDD | 297 | 155 | 49 |  |
| BP | 42 | 21 | 9 |  |
| SZ | 12 | 9 | 4 |  |
| SZA | 22 | 7 | 3 |  |
| **Remission state at baseline n<sup>1</sup>** |  |  |  | **p=.334** |
| Acute | 134 | 84 | 23 |  |
| Partial Remission | 88 | 44 | 17 |  |
| Full Remission/Symptom free | 129 | 53 | 18 |  |
| **Psychiatric comorbidity at baseline n** |  |  |  | **p=.015\*** |
| None | 243 | 102 | 36 |  |
| At least one | 130 | 90 | 29 |  |
| **Recurrence status MDD n** |  |  |  | **p=.099** |
| Single Episode | 117 | 45 | 18 |  |
| Recurrent | 187 | 113 | 31 |  |

*Note: p-Values indicate results for Chi-Square test or ANOVA. Symptom change data were missing for n=16 patients. Stars indicate significance at p<.05. 1) Data were missing for n=42 participants*

Table 6 Cont.: Patients grouped according to symptom changes since the pandemic
| Changes in symptom severity from January to April/May 2020 | Unchanged | Worsened | Improved | difference (p-value) |
| --- | --- | --- | --- | --- |
| **Covid-19 data** |  |  |  |  |
| Number of experienced Covid-19 incidents n (SD) | 2.84 (1.66) | 3.22 (1.81) | 3.12 (1.85) | $p=.095$ |
| Negative Covid-19 impact rating | 5.64 (4.76) | 7.84 (5.56) | 5.63 (5.54) | $p<.001^*$ |
| Positive Covid-19 impact rating | 1.01 (2.33) | 0.63 (1.36) | 1.45 (2.59) | $p=.017^*$ |
| Total Covid-19 Impact | 6.64 (5.12) | 8.47 (5.63) | 7.08 (6.77) | $p=.001^*$ |
| Subjective fear | 12.83 (5.21) | 14.84 (6.09) | 12.28 (5.47) | $p<.001^*$ |
| Subjective isolation | 5.35 (2.18) | 7.11 (2.44) | 5.95 (2.29) | $p<.001^*$ |
| **Experienced Covid-19-related constraints in treatment options n** | 78 | 79 | 19 | $p<.001^*$ |
| Problems obtaining medication (n) | 6 | 12 | 0 | $p = .003^*$ |
| Problems obtaining social psychiatric aid (n) | 19 | 18 | 8 | $p = .040$ |
| Changes in therapy (n) | 55 | 67 | 16 | $p<.001^*$ |
| Other changes (n) | 15 | 16 | 4 | $p = .103$ |

*Note: p-Values indicate results for Chi-Square test or ANOVA. Symptom change data were missing for n=16 patients. Stars indicate significance at $p<.05$ .*

**Figure 1.**
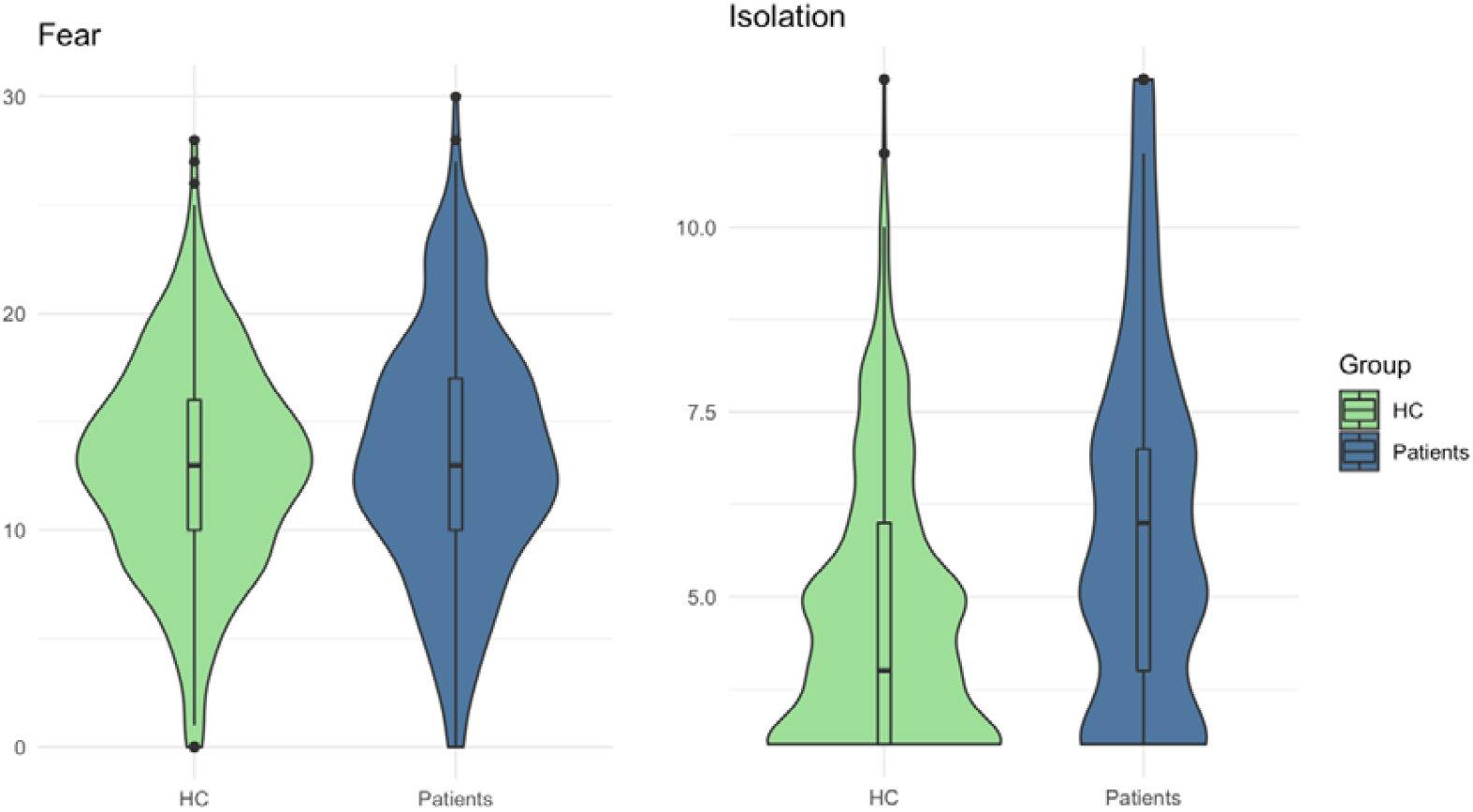
Subjective Covid-19 distress during lockdown in patients and HC. *Note:* Higher values indicate higher subjective isolation/fear. Patients and healthy controls (HC) do not differ significantly in subjective fear (range: 0-30), but in subjective isolation (range: 3-12).

**Figure 2.**
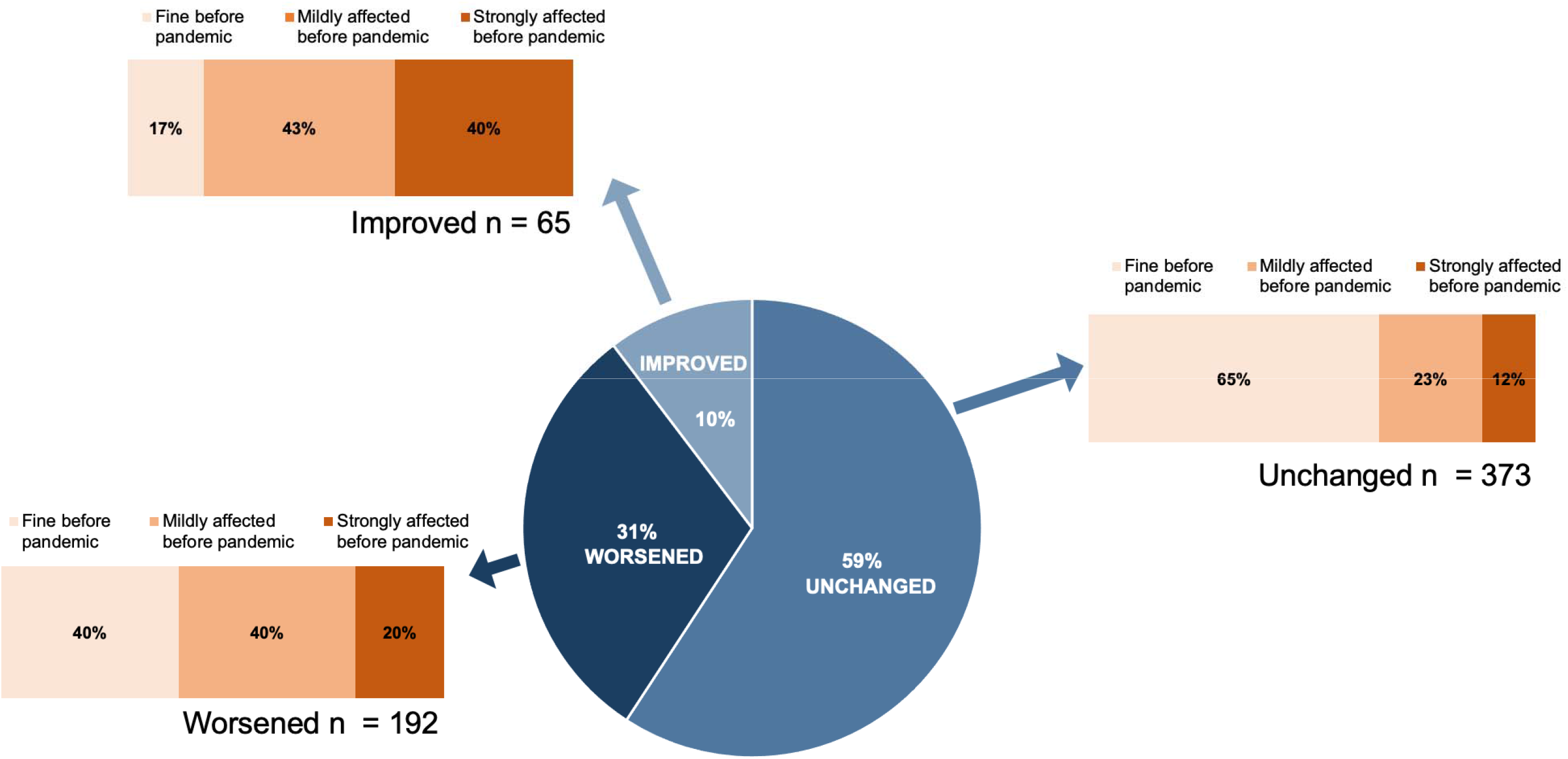
Symptom changes in patients since beginning of pandemic and pre-pandemic mood state.

Table 6 lists differences in the three symptom change groups. Patients reporting worsened symptoms since pandemic onset did not statistically differ from those with improved or unchanged symptoms with regard to the baseline variables: diagnosis (χ^2^ (6) = 3.42, *p*=.755), treatment state (χ^2^(4) = 4.57, *p*=.334), or recurrence status of MDD (χ^2^(2) = 4.62, *p*=.099). They did not differ in number of Covid-19-related incidents during the pandemic (*p*=.095), however, they differed significantly in their negative Covid-19 impact rating (*F*(2,627)=12.53,*p*<.001,η^2^=.038), positive Covid-19 impact rating (*F*(2,627)=4.10,*p*=.017,η^2^=.013), and their total Covid-19 impact rating (*F*(2,627)=7.16,*p*=.001,η^2^=.022).

The groups also differed significantly in both measures of subjective Covid-19 distress, i.e., subjective fear (*F*(3,642)=6.58, *p*<.001), and subjective isolation: *F*(2,629)=38.26, *p*<.001. (See Table 6).

### Regression models predicting Covid-19 distress using baseline data

*Subjective fear* was statistically significantly predicted by the model, *F*(16,1208)=4.71, *p*<.001, *R*^2^=.06 (adj.*R*^2^=.05), indicative of a weak goodness-of-fit (Cohen, 1988). Higher trait anxiety (*β*=.185, *p*_adj_=.006), higher Covid-19 impact (*β*=.130, *p*_adj_<.001), lower conscientiousness (*β*=.111, *p*_adj_=.004), and sex (*β*=.078, *p*_adj_=.020) significantly predicted higher subjective Covid-19 fear. The other variables did not predict subjective fear (i.e., diagnosis, age, site, childhood maltreatment, social support, openness, extraversion, agreeableness, neuroticism, resilience, IQ, familial risk; adj. *p*s >.05) (See Supplement Table 1).

As *subjective isolation* differed statistically in HC and patients, we analysed multiple regression models for HC and patients independently. In HC, subjective isolation was significantly predicted by the model: *F*(15,582)=8.17, *R*^2^=.17 (adj.*R*^2^=.15), indicative of a moderate goodness-of-fit (Cohen, 1988). Lower social support (*β*=-.194, *p*_adj_<.001), sex (*β*=.161, *p*_adj_<.001), lower age (*β*=-.177, *p*_adj_<.001), and higher Covid-19 impact (*β*=.131, *p*_adj_=.002) predicted higher subjective isolation in HC. The other variables did not predict subjective isolation in HC (i.e., site, childhood maltreatment, trait anxiety, Big Five, resilience, IQ, familial risk, all adj. *p*s >.05), see Supplement (Table 2).

In patients, subjective isolation was significantly predicted by the model, *F*(15,610)=9.99, *p<*.001, *R*^2^=.20 (adj.*R*^2^=.18), indicative of a moderate goodness-of-fit (Cohen, 1988). In patients, higher Covid-19 impact (*β*=-.114, *p*_adj_=.006), and lower social support (*β*=-.269, *p*_adj_<.001) predicted higher subjective isolation. The other variables (i.e., age, sex, site, childhood maltreatment, trait anxiety, Big Five, resilience, IQ, and familial risk; adj. *p*s >.05) did not predict subjective isolation in patients. Summaries of the entire models can be found in the Supplement (Table 1-3).

Using the identified variables for Covid-19 distress (i.e., trait anxiety, Covid-19 impact, conscientiousness, sex, and social support), we re-examined scores in improved, unchanged, and worsened patient groups. We found significantly different sex distribution in these groups, and significantly higher trait anxiety in the worsened group (See Table 6). Patient symptom change groups did neither differ significantly in conscientiousness and social support, nor in Covid-19 impact (see Table 6 and Table 4). Table 4 in the Supplement lists additional patient group characterization by diagnosis.

## Discussion

Based on a large longitudinal, transdiagnostic, bi-centre cohort of patients with mental disorders and healthy subjects, our study provides evidence for a disproportionately stronger subjective isolation effect in psychiatric patients, compared to HC, despite similar number of Covid-19-related events and similar impact rating in HC and patients.

We identified several variables (social support, trait anxiety, conscientiousness, sex, age) associated with Covid-19 distress in HC and patients. These findings point to prolonged stress associated with social isolation. We identified risk populations within the general population and those suffering from mental disorders, for which targeted interventions are warranted.

First, our findings demonstrate that patients are at particular risk for higher subjective isolation when exposed to social distancing schemes. Possible reasons for this might include general or non-specific elevation of stress in a society, in addition to lack of resilience, resources, and reduced availability of psychiatric support. Such effects have been demonstrated in both adults and adolescents, usually not in cohorts that have had previous clinical characterization for pre-Covid disease courses (Loades et al., 2020; Riehm et al., 2020). Our findings align with two meta-analyses identifying psychiatric patients as at-risk for increased mental distress during the pandemic (Luo et al., 2020; Xiong et al., 2020). It is possible that healthy individuals have more resources for emotional/social support at their disposal (as indicated by higher baseline social support) that buffered the negative effects of these incidents and prevented higher feelings of isolation (Hoefnagels, Meesters, & Simenon, 2007). 30.5% of our patients reported worsened symptoms since the pandemic. This group of patients also rated Covid-19-related incidents more negatively, less positively, and reported higher subjective isolation and -fear compared to the other patient groups. Restoring sufficient patient care might be one important aspect in improving symptom load (as problems in this area were reported by 27.9% of all patients). In the general population, more than one third of adults with serious distress during the pandemic listed inability to obtain health care as a contributing factor for increased distress, further highlighting the need to facilitate (physical and mental health) treatment attainment (McGinty, Presskreischer, Anderson, Han, & Barry, 2020).

Second, using baseline data, we were able to predict Covid-19 distress, i.e., subjective fear and isolation. Higher subjective fear was significantly predicted by higher trait anxiety, higher Covid-19 impact, lower conscientiousness, and female sex in HC and psychiatric patients (in order of predictive strength, see Supplement). However, these variables only explained 6% of variance in subjective fear. Other current factors, such as media consumption and catastrophic cognitions about Covid-19 might also influence subjective fear, but were not assessed in this study (Bendau et al., 2020; Rosebrock et al., 2021).

Interestingly, the patient group reporting worsened symptoms since the pandemic also reported significantly higher trait anxiety scores than the unchanged and improved groups. Trait anxiety might not only be associated with subjective fear of Covid-19, but also with worse mental health outcomes during the pandemic in general. This aligns with previous findings reporting an association between depressive symptoms and perceived risk of Covid-19 infection (Kim, Nyengerai, & Mendenhall, 2020).

As subjective isolation differed significantly between HC and patients, we ran two independent multiple regression models. In patients, subjective isolation was predicted by social support and Covid-19 impact. In HC, it was predicted by the same factors as patients, but additionally by age (inversely) and sex.

In line with our findings, female gender has been repeatedly associated with poorer mental health outcomes during the pandemic (Luo et al., 2020; Vindegaard & Benros, 2020; Xiong et al., 2020). In patients, sex did not reach significance after correcting for multiple tests (*p*=.051) but might also contribute to subjective isolation in patients.

Our findings show that besides Covid-19 impact, trait variables such as social support (and additionally age and sex in HC) contribute to subjective isolation during the pandemic, with models accounting for 15% of variance in HC, and 18% of variance in patients. Previous social support seems to be important in feeling connected and less isolated during the pandemic. Our findings align with findings by Gloster et al., (2020), who identified social support as a strong predictor for mental health outcomes during the pandemic.

It is noteworthy that social support data, collected 2-6 years prior were still relevant for subjective isolation. Social isolation during the pandemic has also been associated with poorer life satisfaction, and higher levels of substance use (Clair, Gordon, Kroon, & Reilly, 2021). Current social support during the pandemic was found to be associated with fewer depressive symptoms, with quality of contact being more important than quantity (Sommerlad et al., 2021). Higher social support was also associated with more positive appraisal and higher resilience during the pandemic (Veer et al., 2021). In light of these findings, social support seems to be relevant for both HC and patients in the maintenance of mental health during a pandemic.

In the aftermath of traumatic events, a “pulling together effect” is often described, with an increase in social support and cohesion in the affected population. This effect was shown to buffer negative effects of such traumatic events (Mancini, 2020; Reger et al., 2020). However, with increased levels of social isolation, especially in patients, this effect does not seem to be present during this pandemic. This might be explained by the difference between singular traumatic events (e.g., natural disasters, terrorist attacks) compared to prolonged or even ongoing stressors. The specific characteristics of this pandemic and the very strategies (i.e., social distancing) to ameliorate its adverse effects seem to impede the protective effect of increased social cohesion. As social distancing is used to contain virus spread, we as a society have to act counter-intuitively than we would otherwise in stressful situations: as opposed to finding emotional and physical comfort from our social networks, we are asked to limit social contacts and to engage in socially distanced communication with friends and family. Despite large-scale efforts to minimize social (as opposed to physical) distancing, e.g. through the use of web-based communication, these might not overcome the lack of physical proximity to other humans, which has shown significant benefits to mental and physical health (Jakubiak & Feeney, 2019; Thomas & Kim, 2020). This indicates that individual efforts, and new and creative strategies to keep in touch are not sufficient (or not sufficiently used) to offset feelings of isolation. Social isolation is well known to be detrimental to both physical as well as mental health, and constitutes an important risk factor for MDD (Holt-Lunstad et al., 2015; Qiu et al., 2020). Feelings of isolation might add up over time with detrimental effects to mental health, and generate considerable clinical burden.

To counteract isolation during the pandemic, it seems vital to increase social integration and to immunize isolated individuals against the negative impact of restrictions to contain virus spread. Future policies should therefore address the problem of loneliness and isolation and focus on interventions that target feelings of isolations in population groups already experiencing lesser degrees of social integration, and provide safe options for social interaction during the pandemic. Our results point to social support as an important starting point for targeted interventions and preventions.

### Limitations

Several limitations should be noted. The current study did not include a full diagnostic interview at mid-pandemic follow-up, only one conducted for baseline data collection. More objective measures, including HAMD scores, assessed by trained raters and in-depth interviews would have improved the validity of the findings.

We did not re-assess trait anxiety and social support during the pandemic. It might be argued that subjective fear and subjective isolation are simple expressions of these traits, manifesting irrespectively of the pandemic. However, in all three multiple regression models, Covid-19 impact was a significant predictor for subjective fear and isolation. Covid-19-related fear and isolation ratings seem to be influenced by both baseline trait variables (trait anxiety and social support), but also by Covid-19 impact. The found associations cannot be regarded as causal inferences, however, they might guide future research and hint at possible risk factors.

We asked patients about subjective symptom changes between two time points, between January 2020 before the pandemic, and current symptoms during lockdown. While self-ratings are known to be associated with clinicians’ ratings of severity, it is still possible that current symptom load might have influenced symptom severity assessment of January 2020 (Tondo, Burrai, Scamonatti, Weissenburger, & Rush, 1988).

### Outlook

The Covid-19 pandemic constitutes an unfamiliar challenge to individuals and societies worldwide. With ongoing or intermittent lockdowns set in place, policy makers should acknowledge the mental health impact of social isolation, which might be even more pronounced in the long-term (Daly, Sutin, & Robinson, 2020). Targeted prevention and interventions, especially in those populations at particular risk should be considered (Saltzman, Hansel, & Bordnick, 2020). The lack of such interventions might result in significant and lasting impact on mental health burden, in people previously affected by mental health impairments, as well as those without a prior history of mental illness (Galea, Merchant, & Lurie, 2020).

## Data Availability

Data is available from the corresponding author on reasonable requests.

## Acknowledgments

This work is part of the German multicentre consortium “Neurobiology of Affective Disorders. A translational perspective on brain structure and function“, funded by the German Research Foundation (Deutsche Forschungsgemeinschaft DFG; Forschungsgruppe/Research Unit FOR2107).

We are deeply indebted to all study participants and staff. A list of acknowledgments can be found here: for2107.de/acknowledgements.

## Ethical standards

The authors assert that all procedures contributing to this work comply with the ethical standards of the relevant national and institutional committees on human experimentation and with the Helsinki Declaration of 1975, as revised in 2008.

