## Supplementary Material for "Which traits predict elevated distress during the Covid-19 pandemic? Results from a large, longitudinal cohort study with psychiatric patients and healthy controls"

Table 1: Summary for model predicting Covid-19 fear in whole sample

|  | Non-standardized coefficient | | Standardized coefficient | T | *p* | Collinearity Statistics | | *p*_adj_ |
| --- | --- | --- | --- | --- | --- | --- | --- | --- |
|  | Unstandardized coefficient b | Standard error | Beta (β) |  |  | Tolerance | VIF |  |
| Diagnosis  Age  **Sex**  Site  **Covid-19 impact**  Childhood maltreatment  Social support  **Trait anxiety**  Openness  **Conscientiousness**  Extraversion  Agreebaleness  Neuroticism  Resilience  IQ  Familial risk | -.450 | .408 | -.043 | -1.101 | .271 | .519 | 1.927 | 0.813 |
|  | .018 | .013 | .045 | 1.392 | .164 | .745 | 1.342 | 0.493 |
|  | .862 | .333 | .078 | 2.586 | .010 | .862 | 1.161 | **0.029** |
|  | -.494 | .308 | -.047 | -1.603 | .109 | .922 | 1.084 | 0.327 |
|  | .121 | .026 | .130 | 4.581 | .000 | .973 | 1.028 | **0.000** |
|  | .007 | .013 | .017 | .513 | .608 | .673 | 1.486 | 1.825 |
|  | .379 | .280 | .055 | 1.357 | .175 | .478 | 2.092 | 0.525 |
|  | .071 | .024 | .185 | 2.962 | .003 | .199 | 5.029 | **0.009** |
|  | -.031 | .025 | -.039 | -1.238 | .216 | .779 | 1.283 | 0.648 |
|  | .082 | .026 | .111 | 3.121 | .002 | .620 | 1.613 | **0.006** |
|  | .013 | .028 | .020 | .476 | .634 | .429 | 2.333 | 1.902 |
|  | .007 | .030 | .008 | .237 | .813 | .701 | 1.428 | 2.439 |
|  | -.001 | .030 | -.003 | -.045 | .964 | .221 | 4.532 | 2.893 |
|  | -.012 | .011 | -.059 | -1.099 | .272 | .267 | 3.744 | 0.817 |
|  | .003 | .012 | .008 | .255 | .799 | .766 | 1.306 | 2.397 |
|  | .275 | .337 | .023 | .815 | .415 | .962 | 1.039 | 1.245 |

Dependent Variable: Covid-19 related fear

*Note*: In bold: significant predictors of the model after Bonferroni *p*-adjustment for three tests.

Table 2: Summary for model predicting Covid-19 isolation in healthy controls

|  | Non-standardized coefficient | | Standardized coefficient | T | *p* | Collinearity Statistics | | *p*_adj_ |
| --- | --- | --- | --- | --- | --- | --- | --- | --- |
|  | Unstandardized coefficient b | Standard error | Beta (β) |  |  | Tolerance | VIF |  |
| **Age** | -.023 | .006 | -.177 | -3.945 | .000 | .704 | 1.421 | **0.000** |
| **Sex** | .595 | .152 | .161 | 3.909 | .000 | .841 | 1.188 | **0.000** |
| Site | -.099 | .143 | -.027 | -.692 | .489 | .929 | 1.076 | 1.467 |
| **Covid-19 Impact** | .042 | .013 | .131 | 3.384 | .001 | .953 | 1.049 | **0.002** |
| Childhood maltreatment | .012 | .009 | .055 | 1.277 | .202 | .770 | 1.298 | 0.606 |
| **Social support** | -.662 | .169 | -.194 | -3.925 | .000 | .581 | 1.721 | **0.000** |
| Trait anxiety | .029 | .014 | .137 | 2.174 | .030 | .358 | 2.793 | 0.090 |
| Openness | .014 | .011 | .052 | 1.255 | .210 | .812 | 1.231 | 0.630 |
| Conscientiousness | -.018 | .013 | -.066 | -1.424 | .155 | .663 | 1.507 | 0.465 |
| Extraversion | -.012 | .014 | -.041 | -.877 | .381 | .662 | 1.512 | 1.143 |
| Agreeableness | .011 | .014 | .034 | .771 | .441 | .722 | 1.385 | 1.323 |
| Neuroticism | .002 | .015 | .010 | .153 | .879 | .364 | 2.751 | 2.636 |
| Resilience | .004 | .005 | .043 | .767 | .444 | .458 | 2.185 | 1.331 |
| IQ | .005 | .006 | .038 | .873 | .383 | .739 | 1.353 | 1.149 |
| Familial risk | .073 | .167 | .017 | .437 | .662 | .946 | 1.057 | 1.987 |

Dependent Variable: Covid-19 related isolation

*Note*: In bold: significant predictors of the model after Bonferroni *p*-adjustment for three tests.

Table 3: Summary for model predicting Covid-19 isolation in patients

|  | Non-standardized coefficient | | Standardized coefficient | T | *p* | Collinearity Statistics | | *p*_adj_ |
| --- | --- | --- | --- | --- | --- | --- | --- | --- |
|  | Unstandardized coefficient b | Standard error | Beta (β) |  |  | Tolerance | VIF |  |
| Age | -.014 | .008 | -.078 | -1.850 | .065 | .743 | 1.346 | 0.194 |
| Sex | .481 | .201 | .094 | 2.393 | .017 | .846 | 1.182 | 0.051 |
| Site | -.093 | .181 | -.019 | -.513 | .608 | .937 | 1.067 | 1.823 |
| **Covid-19 Impact** | .047 | .015 | .114 | 3.107 | .002 | .980 | 1.021 | **0.006** |
| Childhood maltreatment | .012 | .006 | .078 | 1.917 | .056 | .803 | 1.245 | 0.167 |
| **Social support** | -.790 | .140 | -.269 | -5.624 | .000 | .576 | 1.738 | **0.000** |
| Trait anxiety | .008 | .012 | .040 | .634 | .527 | .323 | 3.092 | 1.580 |
| Openness | .022 | .015 | .062 | 1.462 | .144 | .736 | 1.359 | 0.432 |
| Conscientiousness | -.028 | .015 | -.082 | -1.843 | .066 | .666 | 1.501 | 0.197 |
| Extraversion | .015 | .016 | .048 | .945 | .345 | .499 | 2.002 | 1.036 |
| Agreeableness | -.012 | .017 | -.030 | -.703 | .482 | .731 | 1.368 | 1.447 |
| Neuroticism | .034 | .017 | .127 | 1.988 | .047 | .320 | 3.123 | 0.142 |
| Resilience | .003 | .006 | .028 | .419 | .675 | .302 | 3.307 | 2.026 |
| IQ | .011 | .007 | .062 | 1.516 | .130 | .774 | 1.291 | 0.390 |
| Familial risk | -.055 | .190 | -.011 | -.287 | .774 | .962 | 1.040 | 2.322 |

Dependent Variable: Covid-19 related isolation

*Note*: In bold: significant predictors of the model after Bonferroni *p*-adjustment for three tests.

Table 4: Patients characterization by diagnosis

|  | MDD (*n*=514) | BP (*n*=74) | SZA (*n*=25) | SZ (*n*=33) | difference  (*p*-value) |
| --- | --- | --- | --- | --- | --- |
| **Baseline Data** | | | | | |
| Sex (f/m) | 352/162 | 43/31 | 15/10 | 17/16 | *p=*.073 |
| Trait anxiety | 52.14 *(12.43)* | 46.36 *(11.34)* | 49.13 *(11.31)* | 46.06 (11.05) | ***p<.001*^a)^** |
| Conscientiousness | 30.20 *(7.31)* | 30.23 *(6.81)* | 29.22 *(7.14)* | 31.16 *(5.64)* | *p=.*805 |
| Social support | 3.83 *(0.84)* | 4.08 *(0.75)* | 3.84 *(0.62)* | 3.98 *(0.67)* | *p=.094* |
| **Remission state at baseline n**  Acute  Partial Remission  Full Remission/Symptom free | 207  132  175 | 31  20  22 | 12  3  4 | 24  2  2 | ***p*<.001** |
| **Psychiatric comorbidity at baseline n**  None  At least one | 316  198 | 42  32 | 12  13 | 22  11 | *p*=.424 |
| **Covid-19 Data** | | | | | |
| Positive rating | 0.97 *(2.23)* | 0.97 *(2.38)* | 0.44 *(1.23)* | 1.15 *(2.25)* | *p=.652* |
| Negative rating | 6.42 *(5.16)* | 6.94 *(6.69)* | 5.48 *(5.50)* | 4.76 *(4.31)* | *p=.209* |
| Total Impact rating | 7.39 *(5.47)* | 7.92 *(7.71)* | 5.92 *(5.62)* | 5.91 *(6.10)* | *p=.235* |
| Subjective fear | 13.19 *(5.46)* | 13.76 *(6.24)* | 14.08 *(5.58)* | 15.06 *(6.14)* | *p=.235* |
| Subjective isolation | 5.92 (2.40) | 5.78 (2.18) | 6.60 (2.92) | 6.03 (2.66) | *p=.519* |

*Note: p-Values indicate results for Chi-Square test or ANOVA. Numbers in bold indicate significance at p<.05. 1)*

a) MDD > BP. Bonferroni corrected
